# Holder-Optimized Elastography Reveals a Reproducible Pressure-Sensitive High-Velocity Area Phenomenon in Breast Cancer–Related Lymphedema

**DOI:** 10.64898/2026.02.25.26344759

**Authors:** Zheng-Yu Hoe, Ruei-Sian Ding, Chen-Pin Chou, Chin Hu, Chao-Hsien Lee, Yen-Dun Tzeng, Cheng-Tang Pan, Ming-Chan Lee, Eric Kin-Lap Lee

## Abstract

**Background:** Breast cancer–related lymphedema (BCRL) requires repeated assessment, but lymphoscintigraphy is unsuitable for such short-interval monitoring. Conventional superficial shear-wave elastography (SWE) is limited by pressure-related acquisition artifacts that may suppress diagnostically relevant signal.

**Purpose:** To determine whether Holder-Optimized Elastography (HOE) enables reproducible visualization of a pressure-sensitive high-velocity area (HVA) phenomenon not reliably demonstrable with conventional handheld SWE and whether visual HVA burden is associated with lymphoscintigraphy-defined obstruction severity.

**Materials and Methods:** In this prospective, single-center observational study, Substudy 1 (go/no-go feasibility; 15 women with BCRL; mean age, 61.6 years ± 11.8 [SD]; with HVA-positive cases selected by design) compared paired HOE and handheld ARFI-SWE acquisitions at 240 matched anatomic sites. Substudy 2 (cross-sectional clinical association; 110 women; mean age, 60.7 years ± 10.8 [SD]) evaluated visually assessed HOE-based HVA burden against lymphoscintigraphy-defined obstruction severity using the Taiwan Lymphoscintigraphy Staging (TLS) system.

**Results:** In Substudy 1, HVAs were visible at 125 of 240 HOE sites (52.1%) versus 17 of 240 handheld sites (7.1%); 108 sites were positive only on HOE and none only on handheld imaging (exact McNemar test, *P* < .001). In Substudy 2, inter-reader agreement was acceptable to good for image-level HVA presence (κ = 0.718) and count (ICC(2,1) = 0.756). Global Mean (GM) yielded the highest area under the ROC curve (AUC) for any obstruction versus no obstruction (AUC, 0.837; 95% CI [0.755, 0.920]); Joint-Excluded Mean Difference (JEΔM) yielded the highest AUC for total versus partial obstruction (AUC, 0.865; 95% CI [0.730, 1.000]). Total-versus-partial estimates should be interpreted cautiously because the total-obstruction group comprised 12 participants.

**Conclusion:** HOE enables reproducible visualization of a pressure-sensitive subcutaneous HVA phenomenon not reliably demonstrated with conventional handheld SWE. Visual HVA burden provides clinically meaningful, though incomplete, information regarding lymphatic obstruction severity in BCRL, supporting HOE as a low-pressure acquisition framework for future quantitative SWE assessment.

**Summary Statement:** Holder-Optimized Elastography reveals a reproducible pressure-sensitive subcutaneous high-velocity-area phenomenon in breast cancer–related lymphedema and yields clinically meaningful, though incomplete, information regarding lymphatic obstruction severity.

**Key Results:**

- In a go/no-go feasibility substudy of HVA-positive cases, visible high-velocity areas were present at 125 of 240 HOE-acquired sites (52.1%) versus 17 of 240 conventional handheld sites (7.1%); 108 sites were exclusively positive with HOE (*P* < .001).
- Inter-reader agreement for image-level HVA assessment was κ = 0.718 for HVA presence and ICC(2,1) = 0.756 for HVA count.
- Global Mean yielded the highest AUC for any obstruction versus no obstruction (0.837), and Joint-Excluded Mean Difference for total versus partial obstruction (0.865; total-obstruction group n = 12; 95% CI [0.730, 1.000]).

## Introduction

Breast cancer–related lymphedema (BCRL) is a common and persistent complication of breast cancer treatment which is characterized by the disrupted drainage of protein-rich lymph fluid from lymphatic vessels and impairs function, quality of life, and survivorship (1–4). Lymphoscintigraphy remains the diagnostic reference standard for identifying lymphatic obstruction and characterizing its severity (5–8), but its invasiveness, radiation exposure, and other practical barriers limit its routine repeated use (6), and the method provides neither structural nor biomechanical information. Other imaging approaches provide structural or physiologic information but do not directly address superficial tissue mechanical behavior (4). Therefore, a need remains for a practical, complementary method for repeated evaluation of upper-limb lymphatic impairment.

SWE, including acoustic radiation force impulse (ARFI)–based approaches, offers a noninvasive means of assessing tissue mechanical changes in lymphedema (9–14). Prior studies have produced heterogeneous findings; while some reported diagnostically useful differences or clinically relevant associations between limbs affected and unaffected by BCRL under conventional acquisition conditions (15–17), others did not (18–21). For example, Chan et al. found that SWE can determine if a limb exhibited any level of lymphatic obstruction (“No vs Any”) but was unable to distinguish between partial and total obstruction (16). Probe and gel-pad pressure can alter apparent shear-wave velocity (SWV) distributions in soft tissues (22,23), especially in superficial tissues (24), and coupling medium pressure and operator technique could also limit image acquisition and reproducibility across studies (14). Therefore, it is possible that at least some heterogeneity between reports could be attributed to acquisition artifact. Identifying a reproducible low-pressure SWE method could therefore reopen a diagnostic window that prior handheld studies may have inadvertently suppressed.

To address such shortcomings, we developed Holder-Optimized Elastography (HOE), a pressure-minimized SWE method that stabilizes the transducer above the skin without direct probe contact. Using HOE, we observed a subcutaneous imaging pattern not previously characterized, termed “high-velocity area(s)” (HVA). The present study was performed to determine whether HVA acquisition could be clinically useful in lymphedema diagnosis and monitoring. We conducted two linked analyses: a substudy of HVA acquisition with HOE versus conventional handheld SWE at matched anatomic sites, and another evaluating the reproducibility of visual assessment of HVA and its association with obstruction detection and severity in BCRL. Visual HVA counting served as the primary quantitative readout for this report. In addition, we explored the question of whether HOE-acquired HVA corresponded to physiological phenomena.

## Materials and Methods

### Study Design and Oversight

This single-center, prospective observational study was approved by the institutional review board (REDACTED FOR PREPRINT), and all participants provided written informed consent. The overall study program was comprised of two linked substudies with a series of case studies in between, all conducted under the same IRB-approved SWE imaging program:

*Substudy 1 (Go/No-Go Feasibility):* This HVA-positive substudy was conducted to test acquisition-dependent visibility at matched sites (rather than clinical prevalence or diagnostic performance).

*Corroborative case studies:* Three cases were included to assess whether HVAs corresponded to physiologic tissue findings rather than artifacts of extremely low-pressure imaging.

*Substudy 2 (Cross-Sectional Clinical Association):* This cross-sectional clinical association substudy evaluated whether visually-assessed HVA burden was reproducible and associated with lymphoscintigraphy-defined obstruction severity in BCRL.

#### Participants

*Substudy 1 (Go/No-Go Feasibility):* Between October 2022 and October 2025, women with BCRL (ISL stage II or higher (25)) were recruited (Fig 1). The sole substudy-specific exclusion criterion was absence of visible HVA on HOE-acquired SWE images. Accordingly, the results from this substudy quantify acquisition dependence within an HVA-enriched sample rather than unbiased clinical prevalence, sensitivity, or specificity.

**Figure 1.**
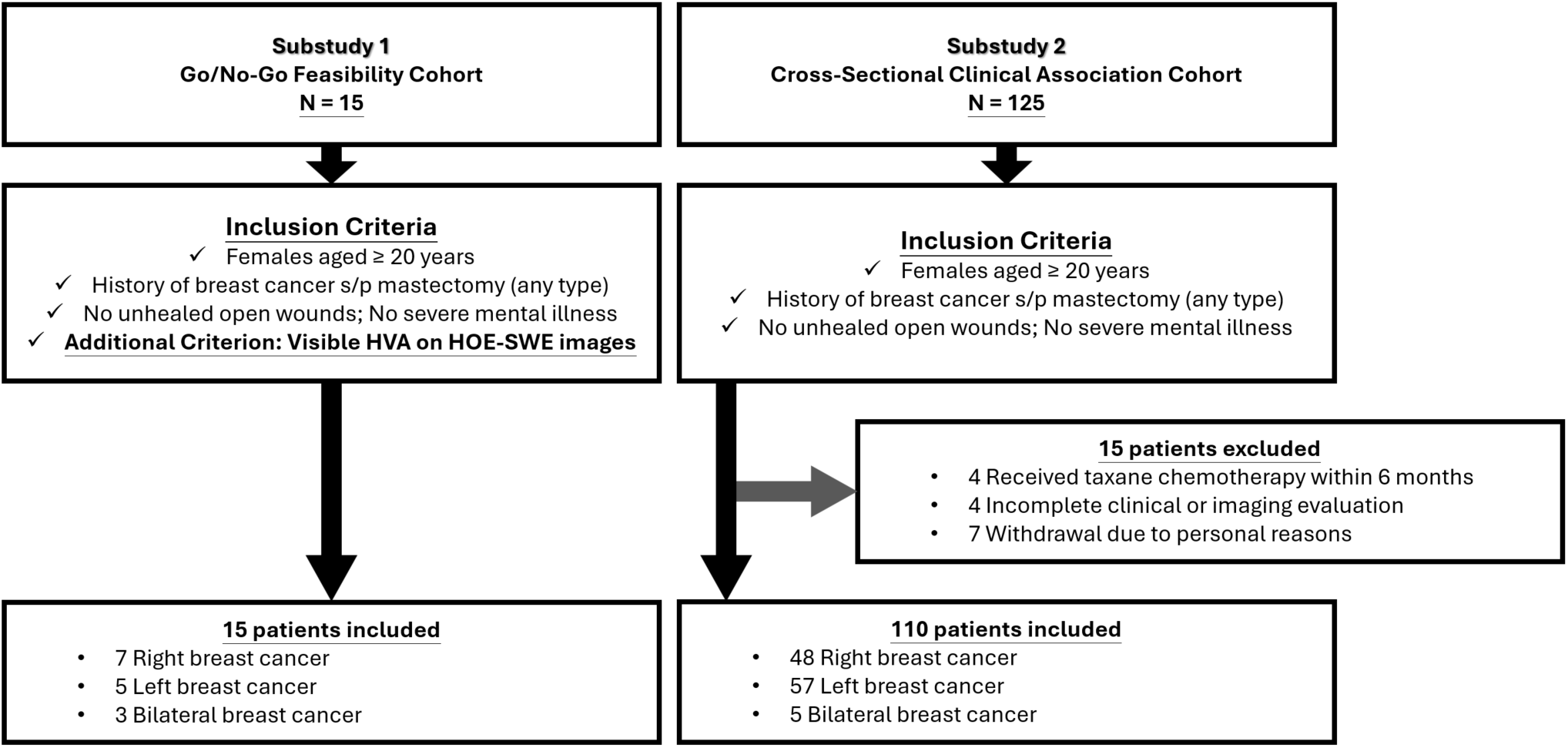
Participant flow diagram for the 2 linked HOE analyses.

*Case studies:* Three case studies are included in this manuscript. Each was the first participant in a distinct clinical circumstance who met the relevant procedural indication and created an ethically feasible opportunity for the next corroborative step. The first participant presented with total obstruction and extensive HVA who agreed to MRI; the second participant’s planned surgical excision overlapped an HVA-localized region and therefore allowed postoperative pathologic correlation without additional tissue sampling; and the final participant underwent lymphaticovenous anastomosis (LVA), allowing preoperative HVA localization to be tested against operative lymphatic identification.

*Substudy 2 (Cross-Sectional Clinical Association):* Between October 2022 and July 2024, women with a history of breast surgery were recruited (Fig 1). Eligibility required age 20 years or older. The exclusion criteria included severe cognitive or emotional disorders, open wounds, bilateral upper-limb swelling from non-BCRL causes, taxane chemotherapy within 6 months, cellulitis within 6 weeks, unreadable lymphoscintigraphy, or incomplete assessments. Participants without lymphatic obstruction were considered postoperative breast cancer controls rather than unrelated healthy volunteers, thereby anchoring comparisons within the same post-treatment clinical setting in which BCRL surveillance is relevant. Clinical covariates including treatment history, lymphedema-related symptom onset, ISL stage, and bilateral upper-limb measurements were recorded at enrollment.

#### HOE Method and Image Acquisition

The core of HOE is mechanical stabilization of the probe above the skin surface, making the coupling gel the only contact interface and preventing anything else from directly pressing on the skin. In this study, the stabilizer was a microphone stand and clip, and a 3D-printed polylactic acid (PLA) coupling gel holder was used to minimize cutaneous loading during ARFI acquisition (Fig 2a and b). The transducer was positioned to contact the coupling gel only, without direct contact with the cutaneous surface or gel holder. This configuration reduced the estimated cutaneous load to approximately 30 g, compared with approximately 160 g during conventional handheld acquisition with a gel standoff pad. As a result, HOE images show more consistent superficial SWV patterns with lower apparent cutaneous SWV and visible scattered “yellow-to-red” high-velocity areas (HVA) in the subcutaneous region (the arrows in Fig 2c), while conventional handheld images show less consistent SWV patterns, relatively increased cutaneous SWV, and no reproducible subcutaneous HVA (Fig 2d). Detailed apparatus specifications, including probe positioning and the rationale for gel-pad omission, are provided in Supplementary Document 1.

**Figure 2.**
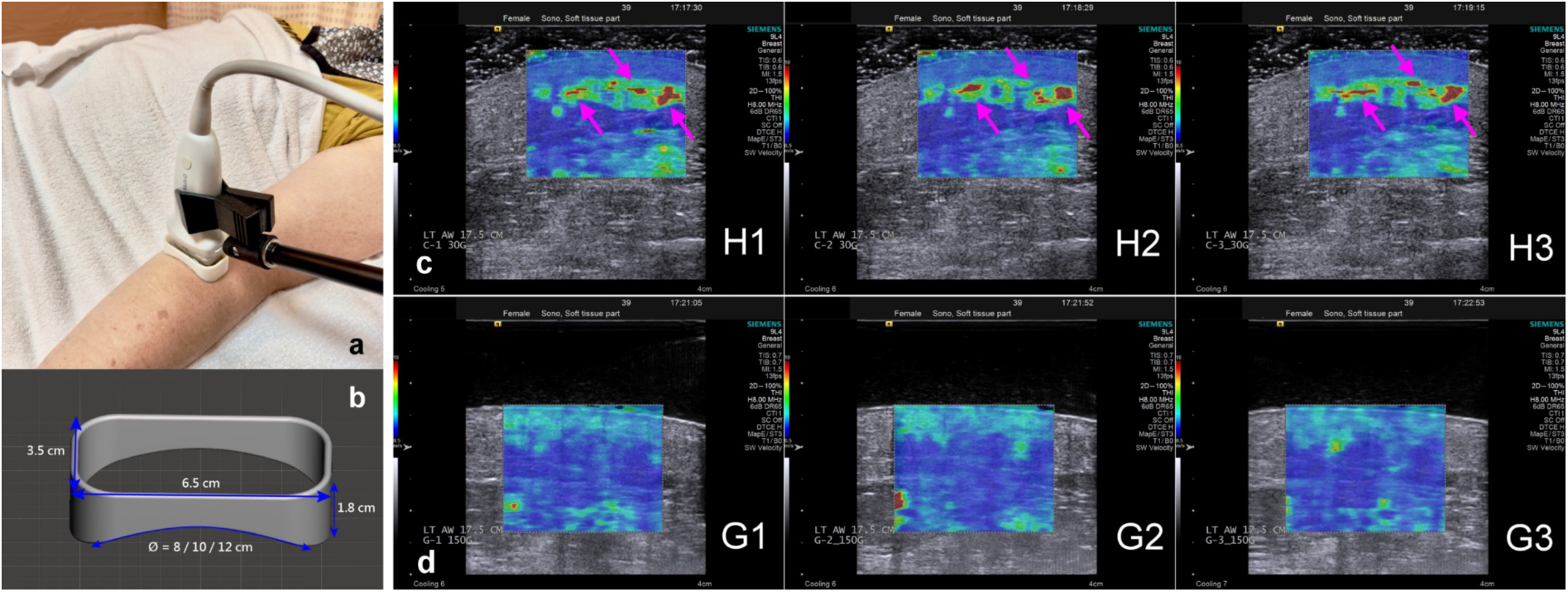
Holder-Optimized Elastography (HOE) setup and representative comparison with conventional handheld acquisition. (a, b) Overview of the HOE setup. (a) The probe is externally stabilized without direct skin contact; the gel receptacle prevents coupling gel from spreading onto the cutis. (b) Gel receptacle design multiple arc radii shown to match varying upper-limb curvatures. (c, d) Comparison of HOE (images H1–H3) versus conventional handheld with gel standoff pad (images G1–G3), acquired from the same lymphedematous region of a woman in her 60s (left arm, 17.5 cm proximal to the wrist crease) at 1-minute intervals.

All examinations used an Acuson S2000 system with Virtual Touch IQ (Siemens Medical Solutions, Mountain View, Calif) and a 9L4 linear array transducer (center frequency, 7.5 MHz; range, 4.0–9.0 MHz). The displayed SWV color scale was fixed at 0.5–10.0 m/s. “SCAN” ultrasound gel (Parker Laboratories, Inc., NJ, USA) was used as the coupling medium.

In both Substudies 1 and 2, participants were positioned supine with palms facing upward. Transverse ARFI images were acquired at 5-cm intervals beginning 2.5 cm proximal to the wrist crease to generate color-coded SWV maps (region of interest depth 2.0 cm, width 2.5 cm, upper boundary aligned with the skin-coupling gel interface).

#### HVA Definition and Reader Process

In this study, an HVA was defined as a visually identifiable subcutaneous region on ARFI color maps with SWV substantially higher than surrounding tissue, appearing as scattered or band-like yellow-to-red foci with relatively clear boundaries (Fig 2c). Under the fixed study-system color scale used herein (0.5–10.0 m/s), such regions typically corresponded to values greater than 7 m/s; this served as a system-specific visual guide rather than a universal cutoff transferable across platforms. Patterns attributable to fascia, muscle interfaces, vessel walls, or other artifactual sources were excluded.

Two physicians experienced in SWE (REDACTED FOR PREPRINT) independently reviewed all images, blinded to participant identity, clinical data, laterality, and lymphoscintigraphy results. Primary interpretation was performed by a radiologist with more than 20 years of ultrasonography experience and familiarity with SWE. Independent verification was performed by a physiatrist with more than 15 years of ultrasonography experience and familiarity with SWE. Each recorded the number of visible HVAs per image; discrepancies were resolved by consensus review.

#### Lymphoscintigraphy and Reference Standard

Planar lymphoscintigraphy of both upper limbs was performed within a 2-week interval of SWE imaging, but not on the same day, because lymphoscintigraphy required radiotracer injection and prolonged same-day ultrasound handling of recently injected limbs would create unnecessary operator radiation exposure without improving interpretation. Two nuclear medicine physicians classified findings by consensus using the TLS system (26) as no obstruction (TLS stage 0), partial obstruction (TLS stages 1–3), or total obstruction (TLS stages 4–6). Detailed protocol and TLS criteria are provided in Supplementary Document 2.

#### Substudy Protocols and HVA-Derived Parameters

*Substudy 1:* Paired HOE and conventional handheld acquisitions were obtained at matched sites. Images were acquired every 5 cm, with a total of eight images obtained per upper limb. The same sequence was repeated on the contralateral, unaffected limb, yielding 16 paired HOE and conventional image sets per patient at identical anatomical levels and under identical positioning and scanning parameters.

*Substudy 2:* Images were acquired using the same 5-cm interval protocol as Substudy 1 but with HOE only (no conventional handheld acquisition). Depending on upper-limb length, up to eight imaging levels were obtained for each limb, and both affected and unaffected limbs were scanned in the same manner. Four direct HVA-count parameters were calculated for each affected limb: Forearm Mean (FM; positions 7.5, 12.5, 17.5 cm from the wrist crease), Joint-Excluded Mean (JEM; positions 7.5, 12.5, 17.5, 27.5, 32.5 cm), Global Mean (GM; mean HVA count across all available positions between 2.5 and 37.5 cm, 6 - 8 positions depending on limb length), and Maximum Value (MAX). Three interlimb difference parameters were then calculated as the affected-minus-unaffected differences of the corresponding direct parameters: FΔM, JEΔM, and GΔM. Variable-site denominators were used without imputation. For participants with bilateral TLS stage 0 findings, the breast cancer surgery side was designated as the affected side for direct-parameter analyses. See Supplementary Document 3 for more details.

### Statistical Analyses

*Substudy 1:* No formal power calculation was performed because Substudy 1 was designed as a go/no-go feasibility analysis of acquisition dependence at matched sites. Visible HVA detection was binarized as present or absent based on whether at least 1 HVA was visible at a matched site. Paired site-level visibility between HOE and conventional handheld ARFI-SWE acquisitions was compared using the exact McNemar test. To account for repeated measurements across multiple sites within the same limb, generalized estimating equation (GEE) logistic regression with a binomial distribution and logit link was performed, with imaging method as the predictor and limb as the clustering unit.

*Substudy 2 Statistical Methods:* Image-level HVA presence agreement was assessed with Cohen’s κ; HVA count (raw counts, and then categorized as 0/1/2/≥3) and derived-parameter agreement were assessed with intraclass correlation coefficients (ICC(2,1) for single measurements, ICC(2,k) for averaged). For affected limbs only, diagnostic performance was evaluated by receiver operating characteristic (ROC) analysis (27) against Taiwan Lymphoscintigraphy Staging (TLS) for 2 comparisons: (a) any obstruction versus no obstruction and (b) total versus partial obstruction. Optimal cutoffs maximized the Youden index (Supplementary Document 3, S3.2 and S3.3).

For both substudies, all analyses were 2-sided; *P* < .05 was considered significant. Statistical analyses were performed using IBM SPSS Statistics for Windows, version 24.0 (IBM Corp, Armonk, NY, USA). Full statistical details are in Supplementary Document 3. Statistical analysis was performed by (REDACTED FOR PREPRINT).

## Results

### Participant Characteristics

Substudy 1 enrolled 15 women with BCRL (mean age, 61.6 years ± 11.8 [SD]; 30 upper limbs; 240 matched sites; Table 1a). Substudy 2 enrolled 110 women after exclusions; TLS classification was no obstruction in 18 (16.4%), partial obstruction in 80 (72.7%), and total obstruction in 12 (10.9%) (Table 1b). The distribution toward partial obstruction reflects the predominant stage in this clinical referral cohort.

**Table 1.** Participant characteristics of two separate substudy cohorts. Table 1a. Go**/No-Go Feasibility Cohort (n = 15)**

| Characteristic | Result |
| --- | --- |
| Age, y | 61.6 ± 11.8 (39.6–80.3) |
| < 40 y | 1 |
| 40–49 y | 1 |
| 50–59 y | 4 |
| 60–69 y | 6 |
| 70–79 y | 1 |
| ≥ 80 y | 2 |
| Body mass index, kg/m<sup>2</sup> | 25.3 ± 3.2 (19.1–29.9) |
| **Surgery type, n(%)** |  |
| Total mastectomy | 11 (73.3) |
| Partial mastectomy | 4 (26.7) |
| **Lymph node surgery, n(%)** |  |
| Axillary lymph node dissection | 9 (60) |
| Sentinel lymph node dissection | 6 (40) |
| **Chemotherapy, n(%)** |  |
| Neoadjuvant | 6 (40) |
| Adjuvant | 9 (60) |
| Without | 0 (0) |
| **Radiotherapy, n(%)** |  |
| Yes | 8 (53.3) |
| No | 7 (46.7) |
| Time from surgery to lymphoedema onset (months) | 60.4 ± 86.9 (–6–311.0) |
| Time from lymphoedema onset to enrollment (months) | 34.1 ± 46.1 (1–167) |
| **ISL clinical stage, n (%)** |  |
| No lymphedema (bilateral TLS stage 0) | 0 (0) |
| Stage 0 (subclinical) | 0 (0) |
| Stage I | 0 (0) |
| Stage II | 12 (80) |
| Stage II (late) | 1 (6.7) |
| Stage III | 2 (13.3) |

**Table 1b.** Cross-Sectional Clinical Association Cohort (n = 110)

| Characteristic | Result |
| --- | --- |
| Age, years | 60.7 ± 10.8 (range, 35–81) |
| <40 y | 1 |
| 40–49 y | 20 |
| 50–59 y | 30 |
| 60–69 y | 30 |
| 70–79 y | 26 |
| ≥80 y | 3 |
| Body mass index, kg/m<sup>2</sup> | 25.2 ± 3.9 (range, 16.8–35.7) |
| **Surgery type, n (%)** |  |
| Total mastectomy | 47 (42.7) |
| Partial mastectomy | 63 (57.3) |
| **Lymph node surgery, n (%)** |  |
| Axillary lymph node dissection | 61 (55.5) |
| Sentinel lymph node dissection | 49 (44.5) |
| **Chemotherapy, n (%)** |  |
| Neoadjuvant | 32 (29.1) |
| Adjuvant | 63 (57.3) |
| None | 15 (13.6) |
| **Radiotherapy, n (%)** |  |
| Yes | 54 (49.1) |
| No | 56 (50.9) |
| Time from surgery to lymphedema onset, mo | 48.6 ± 70.4 (range, 0–311.0) |
| Time from lymphedema onset to enrollment, mo | 23.2 ± 34.9 (range, 0–141.7) |
| **ISL clinical stage, n (%)** |  |
| No lymphedema (bilateral TLS stage 0) | 18 (16.4) |
| Stage 0 (subclinical) | 18 (16.4) |
| Stage I | 14 (12.7) |
| Stage II | 29 (26.3) |
| Stage II (late) | 20 (18.2) |
| Stage III | 11 (10.0) |
| **TLS classification** |  |
| No obstruction | 18 (16.4) |
| Partial obstruction | 80 (72.7) |
| Total obstruction | 12 (10.9) |

Values are mean ± standard deviation (range) or n (%). ISL = International Society of Lymphology; TLS = Taiwan Lymphoscintigraphy Staging. “No lymphedema” indicates postoperative controls with bilateral TLS stage 0. “ISL stage 0” indicates subclinical disease without overt swelling.

### Substudy 1: Acquisition Dependence of HVA Visibility

In the go/no-go feasibility cohort, which included HVA-positive participants by design, HVAs were visible at 125 of 240 HOE-acquired sites (52.1%) versus 17 of 240 conventional handheld sites (7.1%; Fig 3A). At the matched-site level, all 17 handheld-positive sites were also positive on the corresponding HOE acquisition, whereas 108 sites were positive only on HOE and none only on handheld imaging (exact McNemar *P* < .001). Using HOE, HVAs were present in 14 of 15 unaffected limbs but at a markedly lower burden than in affected limbs (unaffected mean = 0.73 HVAs per image vs affected mean = 2.68). GEE analysis showed higher odds of visible HVA detection with HOE than with conventional handheld acquisition (odds ratio, 14.26; 95% CI: [7.42, 27.41]; *P* < .001); this estimate reflects acquisition-dependent visibility within an HVA-enriched sample and should not be interpreted as population-level diagnostic performance. On stratified analysis, HOE-visible HVAs were more frequent on affected than unaffected sites (90 of 120 (75.0%) vs 35 of 120 (29.2%)), whereas handheld visibility remained limited on both sides (16 of 120 (13.3%) vs 1 of 120 (0.8%)); the method-by-limb-status interaction was not significant (*P* = .372; Fig 3B; Table 2).

**Figure 3.**
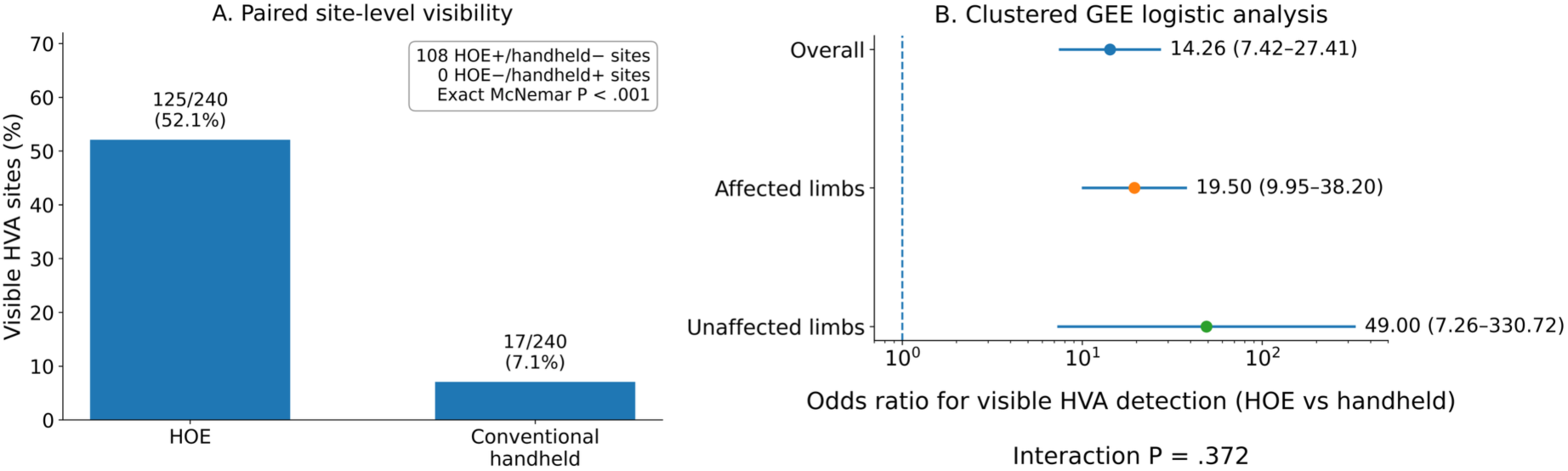
Acquisition-dependent visibility of high-velocity areas in Substudy 1. (A) Paired site-level HVA visibility: 125 of 240 HOE sites versus 17 of 240 conventional handheld sites were positive; 108 sites were positive only on HOE and none only on handheld imaging (exact McNemar P < .001). (B) GEE odds ratios for visible HVA detection with HOE versus conventional handheld imaging, shown overall and stratified by limb status (affected vs unaffected); the method-by-limb-status interaction was not significant (P = .372).

**Table 2.** Stratified site-level HVA visibility results in Substudy 1 according to limb status.

| Limb status | HOE visible HVA / total (%) | Handheld visible HVA / total (%) | # Sites visible in HOE but not in Handheld | # Sites visible in Handheld but not in HOE | Exact McNemar P value | GEE OR (95% CI) | GEE P value |
| --- | --- | --- | --- | --- | --- | --- | --- |
| Overall | 125 / 240<br>(52.1) | 17 / 240<br>(7.1) | 108 | 0 | <.001 | 14.26<br>(7.42–27.41) | <.001 |
| Affected limbs | 90 / 120<br>(75.0) | 16 / 120<br>(13.3) | 74 | 0 | <.001 | 19.50<br>(9.95–38.20) | <.001 |
| Unaffected limbs | 35 / 120<br>(29.2) | 1 / 120<br>(0.8) | 34 | 0 | <.001 | 49.00<br>(7.26–330.72) | <.001 |

Visible HVA = presence of $\geq 1$ HVA at a matched site. Exact McNemar P values compare paired HOE versus conventional handheld acquisition within each stratum. OR = odds ratio estimated by GEE logistic regression with limb as the clustering unit. Method-by-limb-status interaction P = .372.

### Correlative Evidence Supporting the Presence and Nature of HVA

We conducted a number of case studies to establish whether HVA corresponded to actual physiologic phenomena or were artifacts of imaging at extreme low pressures; the results provide corroborative data for a mechanistic explanation of HVA.

In the first case, a woman in her 70s after radical mastectomy presented with total lymphatic obstruction on the right (affected) upper limb. HOE-acquired SWE and MRI T2 imaging demonstrated matching depth relationships and spatial positioning relative to adjacent vascular structures (Fig 4a-d). Specifically, the HVA-localized region corresponded spatially to a superficial fluid-related tubular structure (Fig 4c, d).

**Figure 4.**
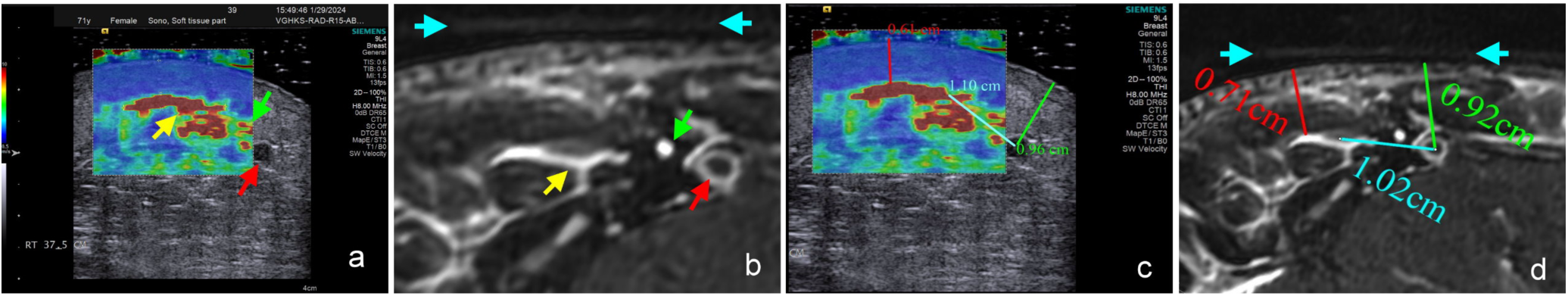
MRI corroboration of HVA-associated regions in a woman in her 70s after radical mastectomy: (a) HOE-acquired SWE of the right upper limb showing a blood vessel (red arrow) and HVA-associated regions (yellow and green arrows). (b) MRI T2 image at the same location with corresponding colored arrows. (c, d) Geographic measurements confirming alignment of superficial structures; blue arrows indicate a fat-filled tube marker identifying probe position and orientation.

In another patient, a woman in her 70s, preoperative HOE was performed prior to modified radical mastectomy. SWE revealed a localized HVA over a breast region that was subsequently excised during surgery (Fig 5a, b), and D2-40 immunostaining of the corresponding specimen demonstrated dilated lymphatic structures in a matching distribution (Fig 5c, d).

**Figure 5.**
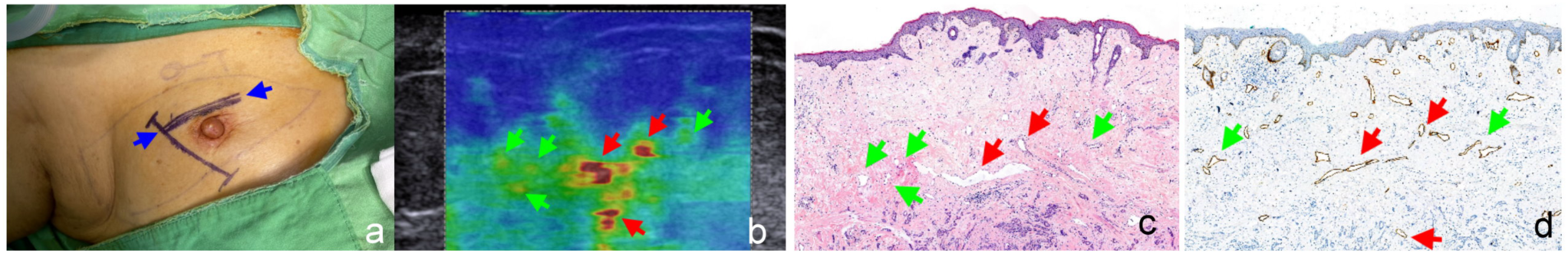
Histopathological corroboration of HVA-associated regions in a woman in her 70s undergoing right modified radical mastectomy. (a) Preoperative planning; blue arrows mark the SWE site within the planned excision field. (b) Preoperative HOE-acquired SWE image; red arrows indicate more conspicuously elevated SWV regions and green arrows indicate adjacent less-marked elevation. (c) Hematoxylin-eosin section. (d) D2-40–positive section showing mildly dilated lymphatic structures (green arrows) and more markedly dilated structures (red arrows) in a distribution corresponding to the HVA-associated region.

In the third case, a woman in her 50s was diagnosed with left-sided breast cancer and left upper-limb lymphedema (TLS P-3) and received lymphaticovenous anastomosis (LVA). Preoperative HOE identified an HVA site (Fig 6a) that was not detected by indocyanine green (ICG) lymphography (Fig 6b). During surgery, a distinct tubular structure was observed precisely beneath the preoperatively marked HVA site (Fig 6c), and intraoperative fluorescence confirmed ICG uptake within the structure (Fig 6d), supporting a lymphatic identity.

**Figure 6.**
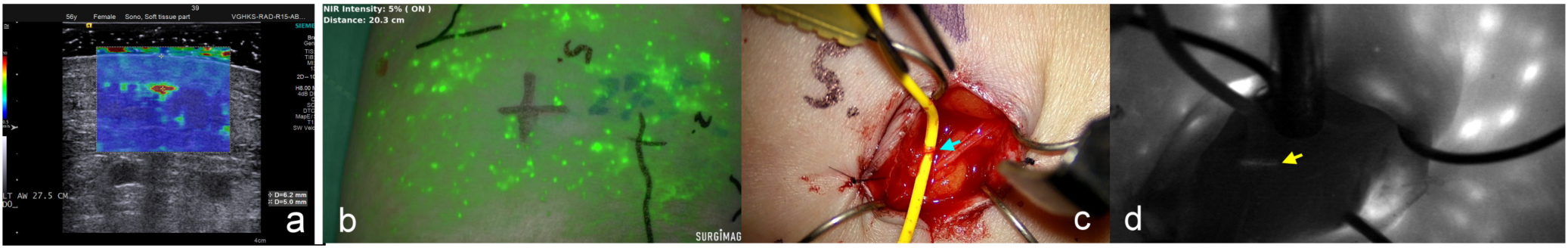
Intraoperative lymphatic identification of an HVA-associated region in a female patient in her 50s who received lymphaticovenous anastomosis. (a) Preoperative HOE identified an HVA at 27.5 cm proximal to the wrist crease, and its skin projection was marked. (b) Preoperative ICG lymphography showed no visible linear pattern near the marked site. (c) Intraoperative view showing a tubular structure beneath the preoperatively marked HVA site (blue arrow). (d) Intraoperative fluorescence confirmed ICG uptake by the structure (yellow arrow).

Additional observations regarding orientation-dependent HVA morphology and real-time pressure-dependent SWV changes during transient compression are provided in Supplementary Document 4 and Supplementary Figures 2 to 4.

### Substudy 2a: Reproducibility of Visual HVA Assessment

1686 evaluable images were acquired with HOE. Cohen’s κ was 0.718 for image-level HVA presence (present vs. absent); ICC(2,1) was 0.756 and ICC(2,k) was 0.861 for image-level HVA count (0, 1, 2, or ≥3). Agreement for derived patient-level parameters was similarly acceptable to good: ICC(2,1) ranged from 0.717 (MAX) to 0.844 (JEM) for direct parameters and from 0.788 (FΔM) to 0.832 (JEΔM) for difference parameters (Table 3).

**Table 3.**
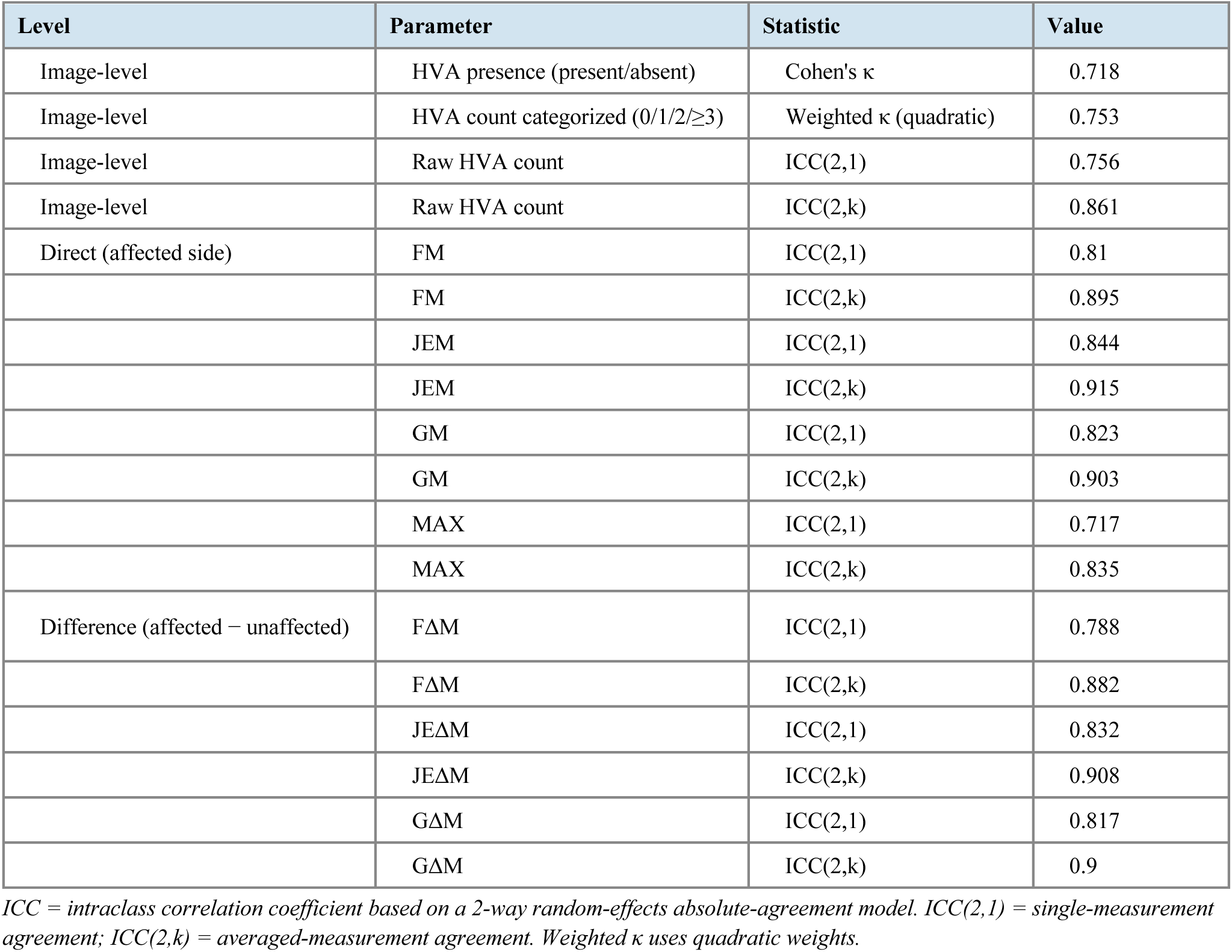
Reader reproducibility for image-level and patient-level visual HVA assessment.

| Level | Parameter | Statistic | Value |
| --- | --- | --- | --- |
| Image-level | HVA presence (present/absent) | Cohen's $\kappa$ | 0.718 |
| Image-level | HVA count categorized (0/1/2/$\geq 3$) | Weighted $\kappa$ (quadratic) | 0.753 |
| Image-level | Raw HVA count | ICC(2,1) | 0.756 |
| Image-level | Raw HVA count | ICC(2,k) | 0.861 |
| Direct (affected side) | FM | ICC(2,1) | 0.81 |
|  | FM | ICC(2,k) | 0.895 |
|  | JEM | ICC(2,1) | 0.844 |
|  | JEM | ICC(2,k) | 0.915 |
|  | GM | ICC(2,1) | 0.823 |
|  | GM | ICC(2,k) | 0.903 |
|  | MAX | ICC(2,1) | 0.717 |
|  | MAX | ICC(2,k) | 0.835 |
| Difference (affected – unaffected) | FΔM | ICC(2,1) | 0.788 |
| | FΔM | ICC(2,k) | 0.882 |
| | JEΔM | ICC(2,1) | 0.832 |
| | JEΔM | ICC(2,k) | 0.908 |
| | GΔM | ICC(2,1) | 0.817 |
| | GΔM | ICC(2,k) | 0.9 |

ICC = intraclass correlation coefficient based on a 2-way random-effects absolute-agreement model. ICC(2,1) = single-measurement agreement; ICC(2,k) = averaged-measurement agreement. Weighted $\kappa$ uses quadratic weights.

### Substudy 2b: HVA Burden and Lymphatic Obstruction Severity

For any obstruction versus no obstruction, all parameters showed significant discrimination (all *P* < .001; Table 4). Among direct affected-limb parameters, GM yielded the highest AUC (0.837; 95% CI [0.755, 0.920]; sensitivity, 0.554; specificity, 1.000); JEM was close behind (AUC, 0.819; 95% CI [0.731, 0.907]; sensitivity, 0.598; specificity, 1.000). Among interlimb difference parameters, GΔM provided the most balanced sensitivity-specificity profile (AUC, 0.818; 95% CI [0.729, 0.906]; sensitivity, 0.783; specificity, 0.778), whereas FΔM and JEΔM prioritized specificity (FΔM: AUC, 0.784; 95% CI [0.685, 0.882]; sensitivity, 0.565; specificity, 1.000; JEΔM: AUC, 0.796; 95% CI [0.702, 0.891]; sensitivity, 0.565; specificity, 1.000).

**Table 4.** Diagnostic performance of visual HVA-derived parameters for lymphoscintigraphy-defined obstruction.

| Comparison | Parameter | AUC (95% CI) | OCV | Sens | Spec | P |
| --- | --- | --- | --- | --- | --- | --- |
| No obstruction vs Any obstruction (n: 18 vs 92) | FM | 0.779 (0.680–0.879) | 1.333 | 0.576 | 1.000 | <.001 |
|  | JEM | 0.819 (0.731–0.907) | 1.200 | 0.598 | 1.000 | <.001 |
|  | GM | 0.837 (0.755–0.920) | 1.286 | 0.554 | 1.000 | <.001 |
|  | MAX | 0.816 (0.727–0.905) | 3.000 | 0.696 | 0.833 | <.001 |
|  | FΔM | 0.784 (0.685–0.882) | 0.667 | 0.565 | 1.000 | <.001 |
|  | JEΔM | 0.796 (0.702–0.891) | 0.800 | 0.565 | 1.000 | <.001 |
|  | GΔM | 0.818 (0.729–0.906) | 0.125 | 0.783 | 0.778 | <.001 |
| Total vs Partial obstruction (n: 12 vs 80)* | FM | 0.822 (0.673–0.972) | 3.000 | 0.833 | 0.725 | <.001 |
|  | JEM | 0.839 (0.695–0.983) | 1.600 | 1.000 | 0.575 | <.001 |
|  | GM | 0.846 (0.704–0.988) | 2.714 | 0.750 | 0.825 | <.001 |
|  | MAX | 0.823 (0.674–0.973) | 6.000 | 0.750 | 0.725 | <.001 |
|  | FΔM | 0.857 (0.719–0.995) | 3.000 | 0.833 | 0.787 | <.001 |
|  | JEΔM | 0.865 (0.730–1.000) | 1.400 | 1.000 | 0.637 | <.001 |
|  | GΔM | 0.862 (0.726–0.998) | 2.750 | 0.667 | 0.912 | <.001 |

*Direct parameters (FM, JEM, GM, MAX) were analyzed on affected limbs only; difference parameters (FΔM, JEΔM, GΔM) = affected minus unaffected side. For participants with bilateral TLS stage 0 findings, the breast cancer surgery side was designated as the affected side for direct-parameter analyses. OCV = optimal cutoff value by Youden index. Sens = sensitivity; Spec = specificity. The total-versus-partial comparison was restricted to the 92 participants with any obstruction and did not include the 18 participants with no obstruction. \*Total-obstruction group n = 12; 95% CIs are wide and should be interpreted cautiously.*

Visual HVA burden increased progressively with obstruction severity. For total versus partial obstruction, all parameters again discriminated significantly; however, the total-obstruction group comprised only 12 participants, and 95% CIs are correspondingly wide. JEΔM yielded the highest AUC (0.865; 95% CI [0.730, 1.000]; sensitivity, 1.000; specificity, 0.637), whereas GΔM yielded the highest specificity (AUC, 0.862; 95% CI [0.726, 0.998]; sensitivity, 0.667; specificity, 0.912) (Table 4). Full ROC results and robustness analyses are provided in Supplementary Document 5, Supplementary Tables 1 and 2.

Because most TLS stage 0 limbs in the cohort were contralateral limbs from participants with unilateral obstruction, we evaluated whether these differed materially from limbs of participants with bilateral TLS stage 0 findings. No material differences were observed for any direct HVA-derived parameter, supporting their combined use in the primary no-obstruction reference group (Table 5).

**Table 5.** Comparability of bilateral-normal and contralateral-normal TLS stage 0 limbs for direct HVA-derived parameters.

| Parameter | Bilateral-normal TLS stage 0 median | Contralateral-normal TLS stage 0 median | Clustered difference | 95% CI | P value | Source-group AUC |
| --- | --- | --- | --- | --- | --- | --- |
| FM | 0.00 | 0.00 | -0.010 | -0.185 to 0.164 | 0.909 | 0.481 |
| JEM | 0.10 | 0.20 | 0.040 | -0.148 to 0.228 | 0.677 | 0.541 |
| GM | 0.25 | 0.25 | 0.024 | -0.172 to 0.220 | 0.808 | 0.508 |
| MAX | 1.00 | 1.00 | 0.147 | -0.529 to 0.823 | 0.670 | 0.507 |

*Bilateral-normal TLS stage 0 = limbs from participants with bilateral TLS stage 0 findings. Contralateral-normal TLS stage 0 = TLS stage 0 limbs from participants with unilateral lymphatic obstruction. Clustered differences estimated accounting for patient-level clustering.*

## Discussion

This study yielded 4 principal findings. First, HOE demonstrated a markedly greater likelihood of visualizing previously unrecognized subcutaneous HVA patterns compared to conventional handheld ARFI, which detected fewer than 1 in 10 sites overall. Secondly, the HVA are not artifacts of a minimal-pressure approach to SWE but rather are associated with real physiological phenomena associated with lymphatic obstruction. Third, visual HVA assessment showed acceptable-to-good reproducibility across both image-level and patient-derived-parameter levels, supporting its applicability as a standardizable readout. Fourth, visual HVA burden showed clinically meaningful, though incomplete, association with lymphoscintigraphy-defined obstruction presence and severity. Overall, we find that HOE serves as a low-pressure acquisition framework that reveals reproducible pressure-sensitive signals. Furthermore, the revealed signals carry clinically relevant information even when assessed with a deliberately simple visual readout.

### HVA Acquisition

The results of Substudy 1 suggested that HVA visualization is dependent on the sensitivity of the mode of acquisition, particularly in subcutaneous tissue. Compared to the traditional handheld SWE, HOE significantly improved the visibility of pressure-sensitive subcutaneous HVA; out of all 240 paired measurement points, the conventional method detected HVA at only 17 measurement points (7.1%). This paired, matched-site data suggest that even modest cutaneous loading, inherent in conventional handheld probe handling (in our case, 160 g with gel pad, measured), can substantially suppress the subcutaneous SWV signal of interest. This could explain why Lee and Cho’s study – which used conventional handheld acquisition with a gel pad – found no significant differences between the SWV of upper limbs with and without BCRL (20). Sanderson et al’s study (which also used handheld acquisition with a standoff pad) showed that localized pressure in the form of the pitting test altered tissue stiffness properties even when removed (21), supporting the pressure-sensitive nature of superficial elastographic assessment. The improved HVA visibility can therefore be attributed, at least in part, to the lower cutaneous load of HOE (30 g of coupling gel). Therefore, HOE appears to do more than just improve handling stability; it may preserve a pressure-sensitive superficial phenomenon that conventional precompression obscures. Accordingly, some prior negative SWE findings in BCRL may reflect methodologic signal suppression rather than a true absence of pathologic abnormality.

A further observation is that HVAs were detectable on unaffected contralateral limbs under HOE (35 of 120 sites, 29.2%), a finding essentially absent on conventional handheld imaging (1 of 120, 0.8%). The large OR for unaffected limbs (49.00) reflects the very low handheld detection rate in this stratum and the wide 95% CI [7.26, 330.72] should be noted. Although markedly less frequent than on affected limbs (75.0% HOE-positive), bilateral detectability raises the possibility that HOE captures physiologically meaningful lymphatic-related SWV phenomena suppressed by conventional precompression. The use of postoperative TLS stage 0 controls rather than unrelated healthy volunteers in Substudy 2 also matters for interpretation: it anchors comparisons within the same post-treatment clinical context as the unaffected contralateral limbs in Substudy 1 and reduces the likelihood that observed differences reflect baseline tissue characteristics unrelated to lymphatic status. Whether subclinical HVA patterns on unaffected limbs carry prognostic significance for future lymphedema development warrants prospective investigation. As the HOE apparatus components used in this study were low-cost and not manufacturer-specific, this may facilitate independent replication and further investigation at centers already performing superficial SWE.

### HVA Correlate With Lymphedema-Related Physiological Phenomena

The three corroborative case studies should be interpreted together as a sequence: from depth-matched imaging, to tissue-level correspondence, and then to direct operative localization. The MRI case established superficial depth- and geometry-matched spatial correspondence with a fluid-related tubular structure; the histopathologic case showed that an HVA-associated region could correspond to dilated D2-40–positive lymphatic structures in resected tissue; and the LVA case showed that a preoperatively marked HVA site could overlie a lymphatic structure identified intraoperatively by ICG uptake. None of these observations alone proves one-to-one identity, but together they argue against display artifact and support a consistent spatial relationship between HVA-localized regions and lymphatic structures or adjacent perilymphatic tissues. This interpretation is further supported by the supplementary mechanistic observations (Supplementary Figures 2 and 3).

The three case studies then provide a mechanistic explanation as to why HOE is better equipped for HVA detection than conventional handheld methods of SWE. It is known that interstitial fluid pressure influences shear-wave behavior (28); however, because fluid-filled spaces themselves do not support shear-wave propagation (11,12), HVA is best interpreted not as direct lymphatic-fluid imaging but as a pressure-sensitive mechanical signal from tissues, interfaces, or remodeled matrix adjacent to fluid-containing lymphatic structures. Chronic lymphatic congestion may also contribute to the presence of HVA through interstitial and adipose tissue remodeling (29). As such, a reasonable interpretation of HVA is a mixed one, in which fluid-related pressure abnormalities and chronic tissue remodeling coexist while HOE reduces enough precompression to preserve the displayed consequence of that abnormal mechanical state. This framework indicates that pressure minimization during acquisition is necessary to preserve the displayed signal, not merely a technical refinement. Separating the relative contributions of fluid-related pressure abnormalities and chronic tissue remodeling to the displayed HVA signal remains an important question for future mechanistic work.

### Reproducibility of HVA Visual Counting

There are few data regarding the reproducibility of visually assessed superficial elastographic patterns, and those assessments are primarily strain elastography (SE) for the evaluation of subcutaneous lesions. Park et al. used a 4-point color scale to grade the SE “hardness” of superficial soft-tissue lesions and found only “moderate” interobserver agreement for the actual 4-point gradation (κ = 0.540); however, when the scores were configured into a binary “soft vs. hard” scale (Grade 1-2 vs. 3-4), the agreement improved to “high” (κ = 0.825) (30). Using the same 4-point color scale but evaluating two images per lesion in their own series, Kim et al. found not only higher interobserver agreement for SE gradation (κ = 0.797 and 0.745 for Session 1 and 2, respectively) but also high intraobserver agreement for each lesion (κ = 0.753 and 0.758 for Readers 1 and 2, respectively) (31). In the present study, our 4-point HVA count system showed higher interobserver agreement than Park et al.’s (weighted κ = 0.753) and comparable agreement for the binary assessment of HVA presence (Cohen’s κ = 0.718). Visual HVA assessment showed acceptable reproducibility at both the image level (κ = 0.718 for HVA presence; ICC(2,1) = 0.756 for HVA count) and patient level (ICC(2,1) ranged from 0.717 to 0.844 for direct parameters and from 0.788 to 0.832 for difference parameters), making our results comparable to both Park and Kim et al.’s, although theirs were of lesion hardness, whereas ours were of discrete counts of focal points. Although only two experienced readers participated in this study, the results support visual HVA counting as a reproducible interim readout; however, its essence remains semi-quantitative. By comparison, Kim et al. found that their quantitative measure called the SE “strain ratio” was more reproducible than their visual color scale in characterizing tissue mechanics.

### Clinical Association of Visual HVA Counting

The clinical association analysis demonstrates that visual HVA burden carries meaningful diagnostic information across the obstruction spectrum. The differing behavior of whole-limb mean parameters and interlimb difference parameters suggests that visual assessment captures two related but nonidentical aspects of disease expression: overall HVA involvement within the affected limb and side-to-side asymmetry. The perfect specificity of GM and JEM for any obstruction indicates that prominent whole-limb HVA burden is a reliable rule-in indicator. Conversely, the modest sensitivity of GM (0.554) confirms that sparse or absent HVA patterns do not exclude early or subtle disease. This pattern of high specificity but modest sensitivity is consistent with the exploratory no-versus-partial obstruction analysis, which showed only moderate discrimination in milder disease (Supplementary Table 3). In such cases, diffuse subcutaneous SWV heterogeneity may not yet form clearly countable discrete foci. HVA patterns showed a severity-related gradient: absent or sparse in limbs without obstruction, focal or clustered in partially obstructed limbs, and broader or more confluent in totally obstructed limbs. This gradient was observed in both whole-limb mean–based and interlimb difference parameters. Boundary ambiguity near tissue interfaces and merging of adjacent foci introduce additional inter-reader uncertainty at intermediate obstruction levels, consistent with the acceptable but imperfect agreement (κ = 0.718; ICC(2,1) = 0.756) and moderate no-versus-partial discrimination (Supplementary Table 3). As a result, broad confluent heterogeneity may be incompletely represented by count-based assessment in advanced disease; therefore, visual HVA counting remains a semi-quantitative readout rather than full-field quantification.

Visual HVA counting served as the primary quantitative readout for this report to test clinical discriminatory capacity independent of image postprocessing and to characterize the inherent limits of manual visual assessment. These observations establish visual HVA counting as a clinically useful rule-in tool and an important interim proof-of-concept readout, but not a stand-alone screening method. HVA counting is ultimately semi-quantitative and does not fully leverage the pixel-level SWV information preserved in each HOE-acquired ARFI image, including HVA area, SWV magnitude, spatial continuity, and diffuse or confluent heterogeneity; these limitations indicate that complementary whole-image quantification is a logical next step. A companion pixel-data analysis of the same HOE image cohort evaluates whether pixel-level SWV quantification can better capture spatial heterogeneity and reduce the sensitivity and dynamic-range limitations of visual HVA counting (32).

## Limitations

This study had several limitations. First, Substudy 1 was restricted to HVA-positive participants by design because the primary question at this stage was acquisition dependence at matched sites rather than prevalence estimation; the reported odds ratio therefore reflects an HVA-enriched sample and does not provide unbiased estimates of HVA prevalence, clinical sensitivity, or specificity. Second, GM and JEM used variable-site denominators, making optimal cutoff values protocol-specific and not directly transferable. Third, the total-obstruction group comprised only 12 participants, yielding wide confidence intervals for severity-discrimination estimates; these should be interpreted with caution. Fourth, reader assessment was performed by only 2 physicians; evaluation by additional readers would further define and validate reproducibility across a broader range of training backgrounds and experience levels. Finally, this was a single-center study limited to upper-limb BCRL; generalizability to lower-limb lymphedema or other etiologies requires prospective validation.

## Conclusion

HOE enables reproducible visualization of a pressure-sensitive subcutaneous HVA phenomenon that conventional handheld SWE does not reliably demonstrate. Visual HVA burden provides clinically meaningful, though incomplete, information regarding the presence and severity of lymphatic obstruction in BCRL. These findings support HOE as a necessary low-pressure acquisition framework for superficial SWE assessment and provide the scientific rationale for subsequent objective whole-image quantification approaches.

## Data Availability

All data produced in the present work are contained in the manuscript and Supplementary Material

## References

1. Gillespie TC, Sayegh HE, Brunelle CL, Daniell KM, Taghian AG. Breast cancer-related lymphedema: risk factors, precautionary measures, and treatments. Gland Surg. 2018;7(4):379.

2. Thomas-MacLean R, Miedema B, Tatemichi SR. Breast cancer-related lymphedema: women’s experiences with an underestimated condition. Can Fam Physician. 2005;51(2):246–7.

3. McEvoy MP, Gomberawalla A, Smith M, Boccardo FM, Holmes D, Djohan R, et al. The prevention and treatment of breast cancer-related lymphedema: a review. Front Oncol. 2022;12:1062472.

4. Pappalardo M, Starnoni M, Franceschini G, Baccarani A, De Santis G. Breast cancer-related lymphedema: recent updates on diagnosis, severity and available treatments. J Pers Med. 2021;11(5):402.

5. Xiong L, Engel H, Gazyakan E, Rahimi M, Hünerbein M, Sun J, et al. Current techniques for lymphatic imaging: state of the art and future perspectives. Eur J Surg Oncol. 2014;40(3):270–6.

6. Moon T, O’Donnell TF, Weycker D, Iafrati M. Lymphoscintigraphy is frequently recommended but seldom used in a “real world setting.” J Vasc Surg Venous Lymphat Disord. 2024;12(2):101738.

7. Pappalardo M, Cheng M. Lymphoscintigraphy for the diagnosis of extremity lymphedema: current controversies regarding protocol, interpretation, and clinical application. J Surg Oncol. 2020;121(1):37–47.

8. Hassanein AH, Maclellan RA, Grant FD, Greene AK. Diagnostic accuracy of lymphoscintigraphy for lymphedema and analysis of false-negative tests. Plast Reconstr Surgery–Global Open. 2017;5(7):e1396.

9. Davis LC, Baumer TG, Bey MJ, Van Holsbeeck M. Clinical utilization of shear wave elastography in the musculoskeletal system. Ultrasonography. 2019;38(1):2–12.

10. Eby SF, Song P, Chen S, Chen Q, Greenleaf JF, An K-N. Validation of shear wave elastography in skeletal muscle. J Biomech. 2013;46(14):2381–7.

11. Taljanovic MS, Gimber LH, Becker GW, Latt LD, Klauser AS, Melville DM, et al. Shear-wave elastography: basic physics and musculoskeletal applications. Radiographics. 2017;37(3):855–70.

12. Bruno C, Minniti S, Bucci A, Pozzi Mucelli R. ARFI: from basic principles to clinical applications in diffuse chronic disease—a review. Insights Imaging. 2016;7(5):735–46.

13. Forte AJ, Huayllani MT, Boczar D, Cinotto G, Ciudad P, Manrique OJ, et al. The basics of ultrasound elastography for diagnosis, assessment, and staging breast cancer-related lymphedema: a systematic review of the literature. Gland Surg. 2020;9(2):589.

14. Nightingale K. Acoustic radiation force impulse (ARFI) imaging: a review. Curr Med Imaging Rev. 2011;7(4):328–39.

15. Polat AV, Ozturk M, Polat AK, Karabacak U, Bekci T, Murat N. Efficacy of Ultrasound and Shear Wave Elastography for the Diagnosis of Breast Cancer-Related Lymphedema. J ultrasound Med Off J Am Inst Ultrasound Med. 2020 Apr;39(4):795–803.

16. Chan W-H, Huang Y-L, Lin C, Lin C-Y, Cheng M-H, Chu S-Y. Acoustic radiation force impulse elastography: Tissue stiffness measurement in limb lymphedema. Radiology. 2018;289(3):759–65.

17. Hayashi N, Yamamoto T, Hayashi A, Yoshimatsu H. Correlation between indocyanine green (ICG) patterns and real-time elastography images in lower extremity lymphedema patients. J Plast Reconstr Aesthetic Surg. 2015;68(11):1592–9.

18. Suehiro K, Nakamura K, Morikage N, Murakami M, Yamashita O, Ueda K, et al. Real-time tissue elastography assessment of skin and subcutaneous tissue strains in legs with lymphedema. J Med Ultrason. 2014;41(3):359–64.

19. Erdogan Iyigun Z, Agacayak F, Ilgun AS, Elbuken Celebi F, Ordu C, Alco G, et al. The role of elastography in diagnosis and staging of breast cancer-related lymphedema. Lymphat Res Biol. 2019;17(3):334–9.

20. Lee DG, Cho JH. Can Tissue Stiffness Measured Using Shear-Wave Elastography Represent Lymphedema in Breast Cancer? Lymphat Res Biol. 2022 Apr;

21. Sanderson J, Tuttle N, Laakso L. Acoustic radiation force impulse elastography assessment of lymphoedema tissue: an insight into tissue stiffness. Cancers (Basel). 2022;14(21):5281.

22. Rominger MB, Kälin P, Mastalerz M, Martini K, Klingmüller V, Sanabria S, et al. Influencing factors of 2D shear wave elastography of the muscle–an ex vivo animal study. Ultrasound Int open. 2018;4(02):E54–60.

23. Alfuraih AM, O’Connor P, Hensor E, Tan AL, Emery P, Wakefield RJ. The effect of unit, depth, and probe load on the reliability of muscle shear wave elastography: Variables affecting reliability of SWE. J Clin Ultrasound. 2018;46(2):108–15.

24. Wang X, Hu Y, Zhu J, Gao J, Chen S, Liu F, et al. Effect of acquisition depth and precompression from probe and couplant on shear wave elastography in soft tissue: an in vitro and in vivo study. Quant Imaging Med Surg. 2020;10(3):754.

25. Lymphology EC of the IS of. The diagnosis and treatment of peripheral lymphedema: 2020 Consensus Document of the International Society of Lymphology. Lymphology. 2020;53(1):3–19.

26. Cheng M-H, Pappalardo M, Lin C, Kuo C-F, Lin C-Y, Chung KC. Validity of the novel Taiwan lymphoscintigraphy staging and correlation of Cheng lymphedema grading for unilateral extremity lymphedema. Ann Surg. 2018;268(3):513–25.

27. Mandrekar JN. Receiver operating characteristic curve in diagnostic test assessment. J Thorac Oncol Off Publ Int Assoc Study Lung Cancer. 2010 Sep;5(9):1315–6.

28. Cihan A, Holko K, Wei L, Vos HJ, Debbaut C, Caenen A, et al. Effect of interstitial fluid pressure on shear wave elastography: an experimental and computational study. Phys Med Biol. 2024;69(7):75001.

29. Tashiro K, Feng J, Wu S, Mashiko T, Kanayama K, Narushima M, et al. Pathological changes of adipose tissue in secondary lymphoedema. Br J Dermatol. 2017;177(1):158–67.

30. Park HJ, Lee SY, Lee SM, Kim WT, Lee S, Ahn KS. Strain elastography features of epidermoid tumours in superficial soft tissue: differences from other benign soft-tissue tumours and malignant tumours. Br J Radiol. 2015;88(1050):20140797.

31. Kim JN, Park HJ, Kim MS, Won SY, Song E, Kim M, et al. The reproducibility of shear wave and strain elastography in epidermal cysts. Ultrasonography. 2022;41(4):698–705.

32. Hoe, Z, Ding, R, Chou, C, Hu, C, Lee, C, Tzeng, Y, Pan, C, Lee, M, & Lee, E. “Pixel-Level Quantification of Breast Cancer–Related Lymphedema Severity from Shear-Wave Elastography: Diagnostic Value of Tissue Stiffness Heterogeneity.” PRE-PRINT FORTHCOMING.

