## Supplemental Documents for "Holder-Optimized Elastography Reveals a Reproducible Pressure-Sensitive High-Velocity Area Phenomenon in Breast Cancer–Related Lymphedema"

### Supplementary Material

*Citation numbers in this Supplementary Material correspond to the reference list in the main manuscript.*

### Supplementary Document 1: Detailed HOE Apparatus and Setup

#### *S1.1 General Concept*

Holder-Optimized Elastography allows for reproducible and ultra-sensitive data acquisition by eliminating as much cutaneous load as possible in a stable manner.

In the present study, the implementation of the HOE method used 2 components to minimize cutaneous loading and operator-dependent variability during ARFI acquisition: an external probe holder and a lightweight gel holder. Note that a gel standoff pad was not employed in HOE to avoid additional cutaneous loading that may affect HVA acquisition (22,23).

#### *S1.2 Probe Holder*

The probe holder secures the ultrasound transducer in a stable position above the target region and eliminates operator-dependent hand pressure. In the present study, a standard microphone stand and microphone clip were used (Supplementary Figure 1a, b), but any configuration that allows stable, hands-free, and vibration-free operation of the transducer may be used. The probe holder allows fine height adjustment to position the transducer at a reproducible distance from the skin surface.

#### *S1.3 Gel Holder*

The gel holder is a lightweight, curved structure that maintains coupling gel at the target location and prevents outflow onto the skin during acquisition. In the present study, the gel holder was fabricated from 3-dimensionally printed polylactic acid (PLA) material, with arc diameters of Ø = 8, 10, and 12 cm to accommodate the curvature of different upper-limb sizes (Supplementary Figure 1c, d).

Preprint

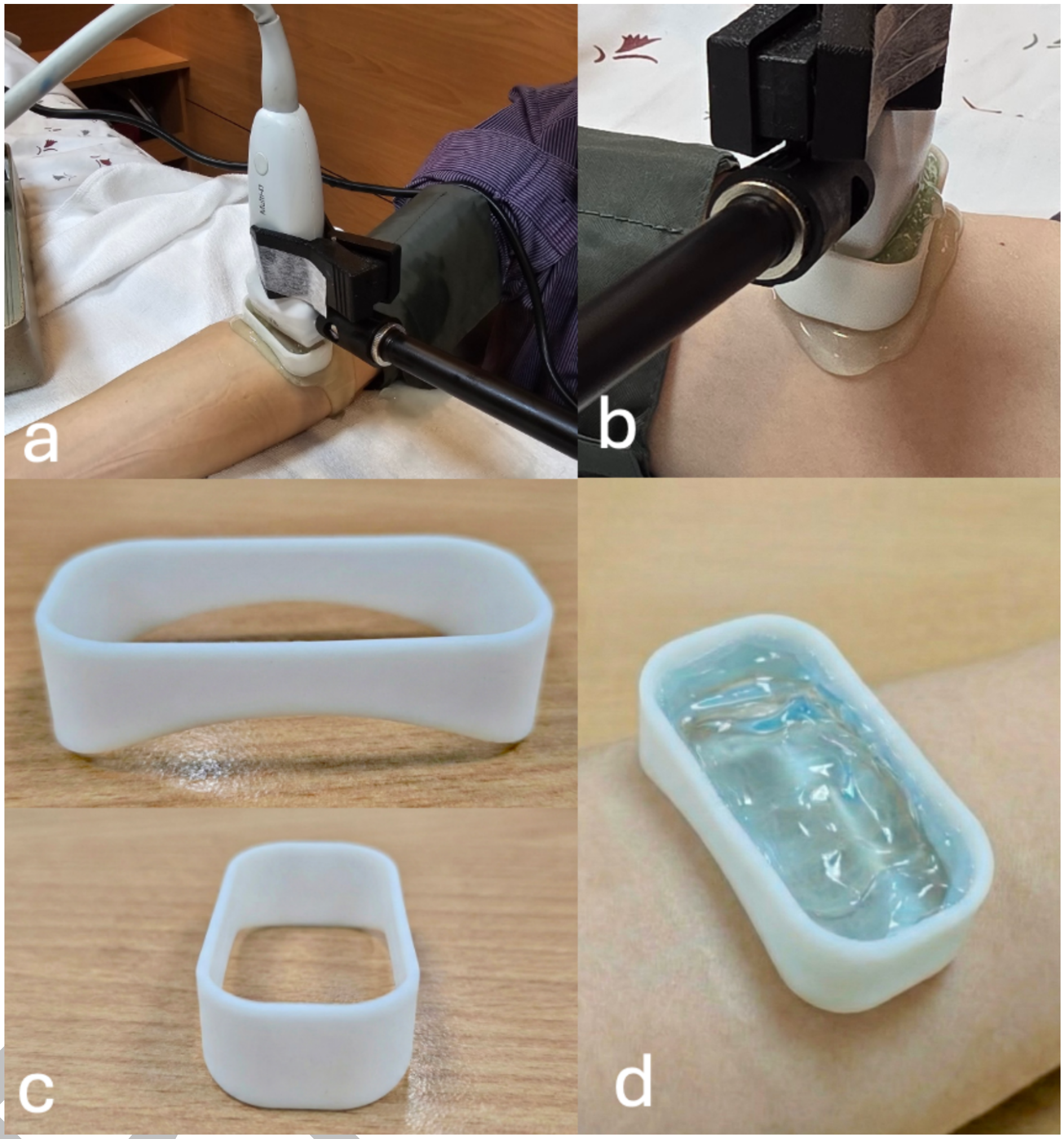


*Supplementary Figure 1. Detailed HOE apparatus and setup. a) Probe holder used in the present study. b) Representative transducer positioning with no direct skin contact. Note that the coupling gel was uncontained and therefore prone to runoff. c) Two views of the coupling gel holder design. d) Gel holder placed over the target region and filled with coupling gel.*

### Supplementary Document 2. Lymphoscintigraphy Protocol and TLS Definitions

#### *S2.1 Imaging Protocol*

Planar lymphoscintigraphy of the bilateral upper extremities was performed at 5–10 minutes and 2–3 hours after subcutaneous injection of technetium-99m phytate into the first web space of each hand. Final interpretation was determined by consensus between 2 nuclear medicine physicians. SWE and lymphoscintigraphy were scheduled within a 2-week interval with no fixed acquisition order, but not on the same day because lymphoscintigraphy required radiotracer injection and prolonged same-day ultrasound handling of recently injected limbs would create unnecessary operator radiation exposure without improving interpretation.

#### *S2.2 TLS Classification Criteria*

Participants were classified according to the Taiwan Lymphoscintigraphy Staging (TLS) system (26) as having no lymphatic obstruction (TLS stage 0), partial obstruction (TLS stages 1–3), or total obstruction (TLS stages 4–6).

#### *S2.3 Use of TLS as the Reference Standard*

TLS served as the reference standard (26) for both primary comparisons: any obstruction (partial or total) versus no obstruction, and total obstruction versus partial obstruction. All TLS interpretations were made blinded to SWE findings.

Preprint

### Supplementary Document 3: Expanded Statistical Methods

All statistical analyses were performed using IBM SPSS Statistics for Windows, version 24.0 (IBM Corp, Armonk, NY, USA).

#### S3.1 HVA-Derived Parameters for Substudy 2

Forearm Mean (FM): mean HVA count at 7.5, 12.5, and 17.5 cm from the wrist crease.

Joint-Excluded Mean (JEM): mean HVA count at positions 7.5, 12.5, 17.5, 27.5, and 32.5 cm, excluding the joint-adjacent levels at 2.5 and 22.5 cm and the most proximal imaged level (typically 37.5 cm).

Global Mean (GM): mean HVA count across all available positions on the limb. This included all positions counted in FM and JEM, plus all previously-excluded positions: 2.5, 7.5, 12.5, 17.5, 22.5, 27.5, 32.5 and 37.5 cm.

Maximum Value (MAX): highest HVA count observed at any available position.

Difference parameters (FΔM, JEΔM, GΔM): affected-side value minus unaffected-side value for FM, JEM, and GM, respectively. Variable-site denominators were used without imputation. For participants with bilateral TLS stage 0 findings, the breast cancer surgery side was designated as the affected side for direct-parameter analyses.

#### S3.2 ROC Analyses

Receiver operating characteristic analyses (27) were performed using Taiwan Lymphoscintigraphy Staging (TLS) as the reference standard (26) for 2 prespecified comparisons: (a) any obstruction versus no obstruction and (b) total obstruction versus partial obstruction. Optimal cutoff values were selected by maximizing the Youden index. If multiple thresholds yielded the same maximum Youden index, the threshold with higher sensitivity was selected; if still tied, the threshold with higher specificity and then the lower cutoff value was selected.

#### S3.3 Supplementary Robustness Analyses

Additional robustness analyses included: a pooled TLS stage 0 sensitivity analysis using both TLS stage 0 limb types as the reference group for direct HVA-derived parameters; and an exploratory no-obstruction versus partial-obstruction analysis to contextualize visual HVA performance in milder disease. None of these exploratory comparisons were prespecified primary analyses.

Preprint

### Supplementary Document 4. Additional Evidence Regarding the Nature of HVAs

#### *S4.1 Pressure Sensitivity and Severity-Related Pattern Changes*

Even minimal pressure applied to the cutaneous surface was sufficient to visibly alter the apparent SWV distribution in the subcutaneous layer. Under the HOE setup, reduced cutaneous loading allowed subcutaneous HVA patterns to remain visually apparent and more clearly delineated. These observations are consistent with prior evidence that superficial tissue mechanical measurements are affected by external loading (14, 24, 30,31) and support a fluid-related mechanical contribution to the HVA phenomenon: fluid-filled spaces do not support shear-wave propagation (11,12), so HVAs most likely represent mechanically stressed perilymphatic tissues rather than direct fluid signal. Representative subcutaneous SWE patterns across obstruction severity levels are shown in Supplementary Figure 2.

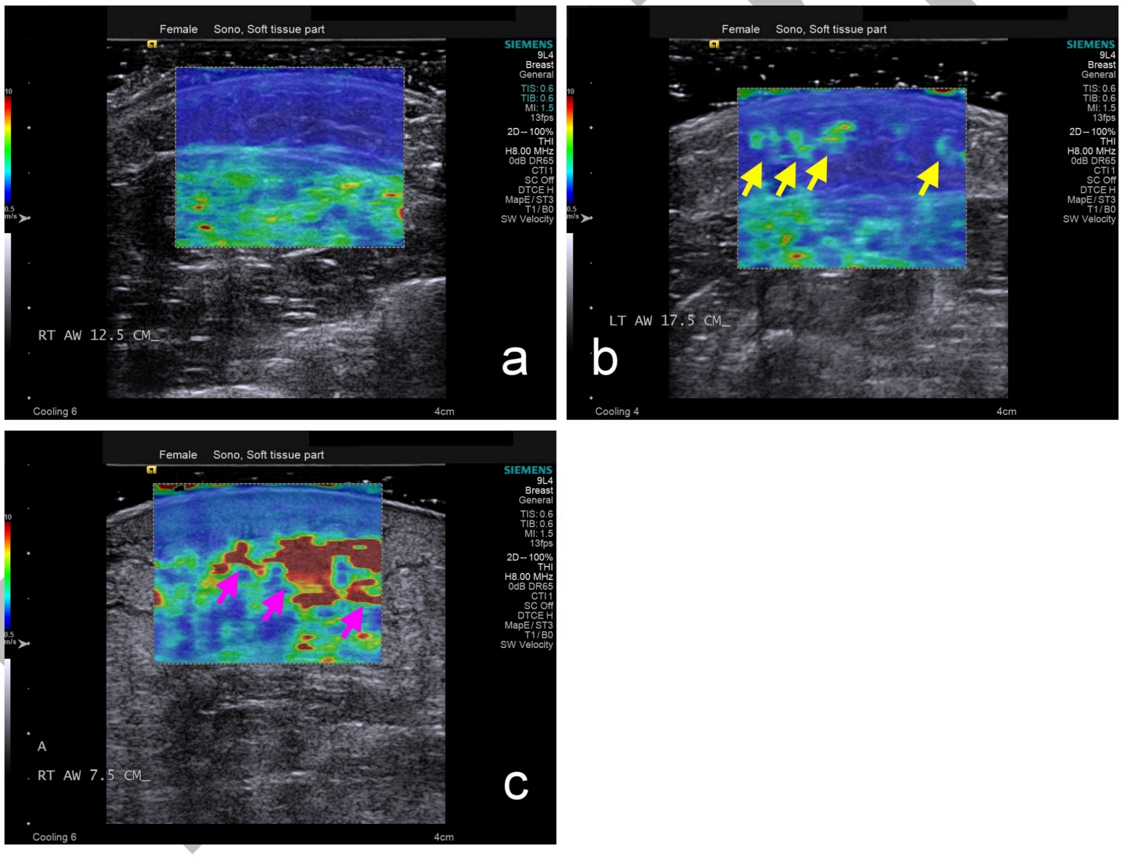


*Supplementary Figure 2. Representative subcutaneous SWE patterns across no, partial, and total obstruction. Subcutaneous shear wave elastography images of the affected limb illustrating visually different patterns across 3 levels of lymphatic obstruction. a) No lymphatic obstruction (postoperative breast cancer control in her 50s without symptoms or clinical manifestations of lymphedema, TLS stage 0): the subcutaneous region demonstrates a relatively homogeneous distribution of apparent shear wave velocity (SWV). b) Partial obstruction (participant in her 50s, TLS stage P2, ISL stage II): focal point-like regions with elevated apparent SWV are observed within the subcutaneous layer (yellow arrows), resulting in a more visually heterogeneous SWV pattern. c) Total obstruction (participant in her 70s, TLS stage P5, ISL stage III): the subcutaneous tissue appears thickened with prominent high-velocity areas (HVAs) (purple arrows), accompanied by a markedly irregular and heterogeneous apparent SWV distribution. The*

*cutaneous layer also appears thickened with visually increased SWV.*

Preprint

### *S4.2 Orientation-Dependent Appearance of HVAs*

The same representative HVA demonstrated distinct morphologic appearances in transverse, oblique, and longitudinal imaging planes. This orientation-dependent appearance is consistent with a tubular or elongated three-dimensional configuration, as expected for lymphatic vessels or perilymphatic fluid collections (Supplementary Figure 3).

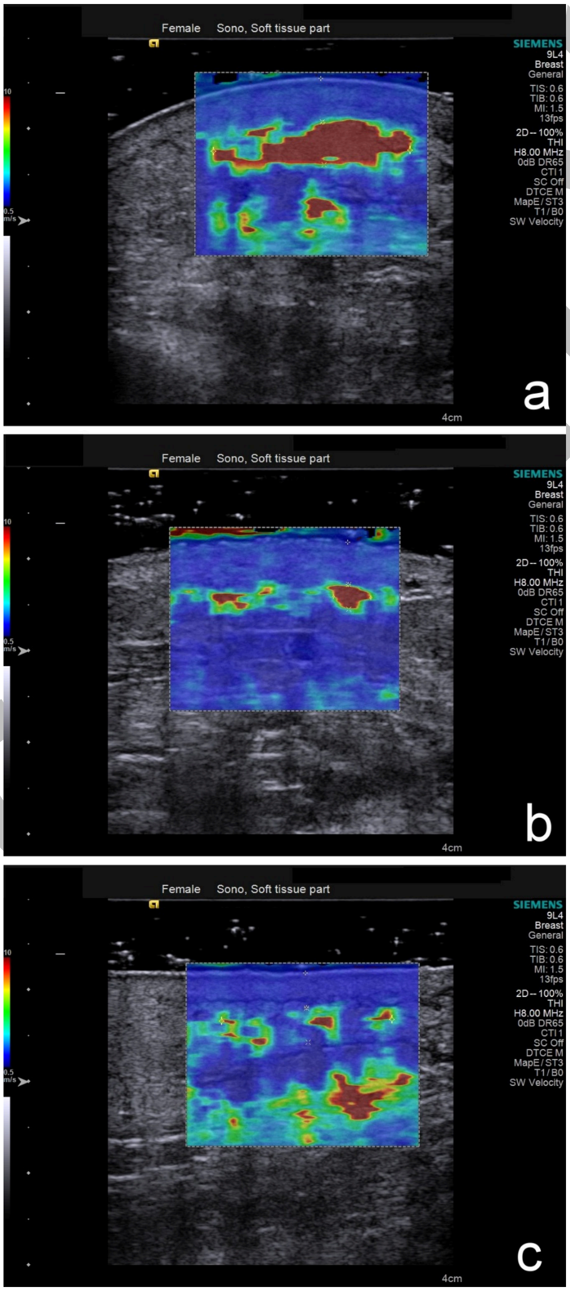


*Supplementary Figure 3. Orientation-dependent appearance of a representative HVA. The same HVA was imaged at the same location under different sectional orientations. a) Transverse section. b) 45-degree oblique section. c) Longitudinal section. The change in apparent shape across different orientations is consistent with a tubular or*

*elongated three-dimensional configuration.*

### S4.3 Real-Time Pressure-Dependent Changes During Transient Compression

Sequential HOE-acquired SWE images obtained before, during, and after transient compression (blood pressure cuff, 40 mmHg for 8 minutes) in a healthy volunteer (female, 20s, no history of cancer or lymphedema) showed that HVA-like regions became more extensive and conspicuous during compression and regressed after release (Supplementary Figure 4). These observations are consistent with prior evidence that interstitial fluid pressure influences shear-wave behavior (28) and that fluid-filled spaces do not support shear-wave propagation (11,12). The pressure sensitivity and reversibility of HVA-like changes support a fluid-related rather than fixed solid-tissue mechanism. This transient compression experiment was intended only to illustrate the pressure-sensitive reversibility of the imaging phenomenon in a controlled setting, not to replicate the pathophysiology of BCRL.

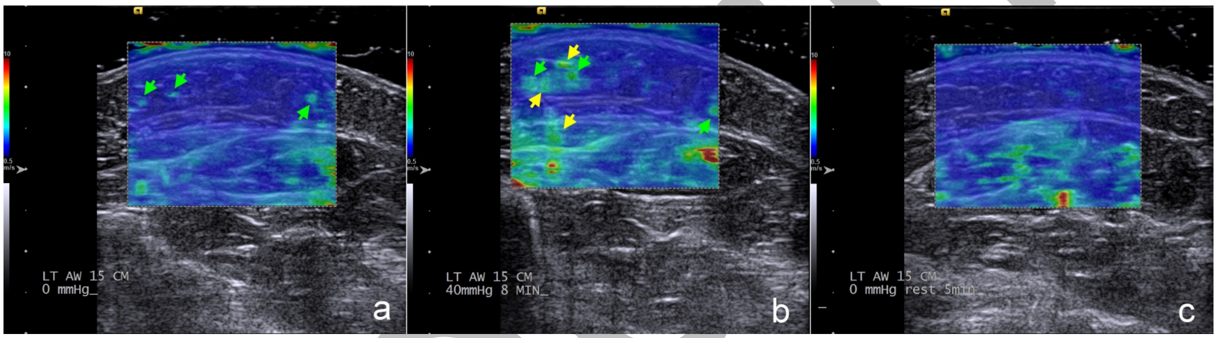


*Supplementary Figure 4. Real-time pressure-dependent changes during transient compression. Real-time visualization of pressure-dependent changes using HOE-acquired SWE images of the forearm (15 cm proximal to the wrist crease) in a healthy female volunteer in her 20s with no history of cancer, systemic disease, or lymphedema, using a blood pressure cuff. a) Before compression, a limited number of focal regions with elevated SWV were present (green arrows). b) During sustained compression (40 mmHg for 8 minutes), pre-existing regions with elevated SWV became more extensive and conspicuous (green arrows), and additional regions with newly elevated SWV emerged (yellow arrows). c) Several minutes after pressure release, the SWE appearance demonstrated gradual regression of the pressure-induced changes, returning toward the pre-compression distribution observed in panel a.*

Preprint

### Supplementary Document 5. Full ROC Results and Robustness Analyses

#### *S5.1 Full ROC Results for the Main Analyses*

Full ROC results corresponding to main manuscript Table 4 are provided in Supplementary Table 1, including sample sizes per comparison. Direct HVA-derived parameters (FM, JEM, GM, MAX) were analyzed on affected limbs only. Difference parameters (FΔM, JEΔM, GΔM) were calculated as affected minus unaffected side.

The total-versus-partial comparison was restricted to the 92 participants with any obstruction and excluded the 18 participants with no obstruction. For participants with bilateral TLS 0 findings, the breast cancer surgery side was designated as the affected side for direct-parameter analyses. GM and JEM used variable denominators (available-site) without imputation.

The 95% CIs are wide and should be interpreted with caution.

Preprint

***Supplementary Table 1. Full ROC results for the main HVA-derived analyses.***

| Comparison | Parameter | AUC (95% CI) | OCV | Sensitivity | Specificity | P-value | n (pos/neg) |
|---|---|---|---|---|---|---|---|
| No vs Any obstruction | FM | 0.779 (0.680–0.879) | 1.333 | 0.576 | 1.000 | <.001 | 92/18 |
| | JEM | 0.819 (0.731–0.907) | 1.200 | 0.598 | 1.000 | <.001 | 92/18 |
| | GM | 0.837 (0.755–0.920) | 1.286 | 0.554 | 1.000 | <.001 | 92/18 |
| | MAX | 0.816 (0.727–0.905) | 3.000 | 0.696 | 0.833 | <.001 | 92/18 |
| | FΔM | 0.784 (0.685–0.882) | 0.667 | 0.565 | 1.000 | <.001 | 92/18 |
| | JEΔM | 0.796 (0.702–0.891) | 0.800 | 0.565 | 1.000 | <.001 | 92/18 |
| | GΔM | 0.818 (0.729–0.906) | 0.125 | 0.783 | 0.778 | <.001 | 92/18 |
| Total vs Partial obstruction* | FM | 0.822 (0.673–0.972) | 3.000 | 0.833 | 0.725 | <.001 | 12/80 |
| | JEM | 0.839 (0.695–0.983) | 1.600 | 1.000 | 0.575 | <.001 | 12/80 |
| | GM | 0.846 (0.704–0.988) | 2.714 | 0.750 | 0.825 | <.001 | 12/80 |
| | MAX | 0.823 (0.674–0.973) | 6.000 | 0.750 | 0.725 | <.001 | 12/80 |
| | FΔM | 0.857 (0.719–0.995) | 3.000 | 0.833 | 0.787 | <.001 | 12/80 |
| | JEΔM | 0.865 (0.730–1.000) | 1.400 | 1.000 | 0.637 | <.001 | 12/80 |
| | GΔM | 0.862 (0.726–0.998) | 2.750 | 0.667 | 0.912 | <.001 | 12/80 |

*OCV = optimal cutoff value by Youden index. *Total-obstruction group n = 12*

### S5.2 Sensitivity Analysis Using Pooled TLS stage 0 Limbs

Because the bilateral-normal and contralateral-normal TLS stage 0 limbs were comparable (Table 5), a sensitivity analysis pooled these groups into a single TLS stage 0 reference for direct HVA-derived parameter comparisons (Supplementary Table 2). Results were consistent with the primary analysis, supporting robustness of the main findings.

***Supplementary Table 2. Sensitivity analysis using pooled TLS stage 0 limbs for direct HVA-derived parameters.***

| Parameter | AUC (95% CI) | OCV | Sensitivity | Specificity | P-value | n (pos/pooled TLS stage 0) |
|---|---|---|---|---|---|---|
| FM | 0.781 (0.716–0.845) | 1.667 | 0.543 | 0.983 | <.001 | 92/116 |
| JEM | 0.798 (0.735–0.860) | 1.200 | 0.598 | 0.940 | <.001 | 92/116 |
| GM | 0.803 (0.741–0.865) | 1.000 | 0.620 | 0.905 | <.001 | 92/116 |
| MAX | 0.780 (0.715–0.845) | 4.000 | 0.630 | 0.888 | <.001 | 92/116 |

Preprint

### *S5.3 Exploratory Analysis: No Obstruction versus Partial Obstruction*

To contextualize the limitation of visual HVA counting in milder disease, an exploratory comparison of no obstruction versus partial obstruction was performed. As shown in Supplementary Table 3, discrimination was only moderate overall and typically favored high specificity over sensitivity, consistent with the hypothesis that early or mild disease produces diffuse subcutaneous SWV heterogeneity without clearly countable HVA margins.

***Supplementary Table 3. Exploratory ROC results for no obstruction versus partial obstruction.***

| Comparison | Parameter | AUC (95% CI) | OCV | Sensitivity | Specificity | P-value | n (pos/neg) |
|---|---|---|---|---|---|---|---|
| No vs Partial obstruction | FM | 0.746 (0.654–0.839) | 1.333 | 0.512 | 1.000 | <.001 | 80/18 |
| | JEM | 0.792 (0.703–0.882) | 1.200 | 0.538 | 1.000 | <.001 | 80/18 |
| | GM | 0.813 (0.721–0.904) | 1.286 | 0.488 | 1.000 | <.001 | 80/18 |
| | MAX | 0.792 (0.691–0.892) | 3.000 | 0.650 | 0.833 | <.001 | 80/18 |
| | FΔM | 0.751 (0.652–0.850) | 0.667 | 0.500 | 1.000 | <.001 | 80/18 |
| | JEΔM | 0.766 (0.671–0.861) | 0.800 | 0.500 | 1.000 | <.001 | 80/18 |
| | GΔM | 0.791 (0.698–0.883) | 0.125 | 0.750 | 0.778 | <.001 | 80/18 |
